# Factors associated with psycho- and pharmacotherapy initiation for common mental disorders in the general population during the COVID-19 pandemic

**DOI:** 10.1101/2025.09.22.25336319

**Authors:** Marie Pouquet, Cécile Vuillermoz, Marion Debin, Camille Davisse-Paturet, Charly Kengne-Kuetche, Clément Turbelin, Thierry Blanchon, Marie Tournier, Olivier Steichen, Nadia Younes

**Author notes:** Author for correspondence: Marie Pouquet, 27 rue de Chaligny 75012 Paris.

## Abstract

**Background:** Health service utilization is influenced by perceived and evaluated health status (need factors), sociodemographic characteristics and health beliefs (predisposing factors), and personal and community resources that facilitate or hinder access to care (enabling factors). No study has simultaneously quantified how these factors directly and indirectly influence psychotherapy and pharmacotherapy initiation for common mental disorders (CMD). The COVID-19 pandemic provides an opportunity to examine these dynamics due to heightened mental health needs and strained healthcare systems.

**Methods:** We conducted a cross-sectional study within the French Grippenet/Covidnet cohort (n=6,944) in April 2022. Data were collected through a voluntary online questionnaire. Participants not using CMD treatments before the pandemic (n=3,297) were included. Weighted structural equation modeling was used to analyze the direct and indirect pathways to psychotherapy and pharmacotherapy initiation.

**Results:** Psychotherapy initiation was directly associated with perceived need (poorer self-rated mental health, negative pandemic impact) and enabling factors (employed, mindfulness/relaxation activities, over-the-counter medication). Pharmacotherapy initiation was directly associated with evaluated need (CMD symptoms, non-psychiatric chronic disease), predisposing (female), and enabling factors (over-the-counter medication). Indirectly, predisposing factors influenced treatment initiation primarily through CMD symptoms (female, younger, lower education, living with adults, adverse life event and recent difficult event), while enabling and pandemic-related factors influenced it through poorer perceived need (loneliness, urban, employed, fewer financial difficulties, less deprived area, pandemic financial decline) and CMD symptoms (loneliness, less sport, most deprived area, symptomatic COVID-19).

**Conclusions:** This study highlights distinct pathways to psychotherapy and pharmacotherapy initiation and provides insights to improve access to each treatment.

## Introduction

Depressive and anxiety disorders are the most common mental disorders (CMDs), with lifetime prevalence estimates of 10.8% for depressive disorders and 28.8% for anxiety disorders [1, 2]. These commonly comorbid disorders substantially impair social, personal, and occupational functioning, with considerable individual and societal economic burdens [3, 4]. Over the past three decades, incidence rates of CMDs have increased, particularly in developed countries [5, 6]. **Potential** contributing factors include financial crises, climate change, geopolitical instability, individualistic societal structures, improved detection, reduced stigma, and, more recently, the COVID-19 pandemic [5–12]. Adequate treatment of CMDs **facilitates** symptom relief and functional recovery [13, 14]. While psychotherapy is the first-line treatment for all levels of severity, pharmacotherapy is recommended in combination with psychotherapy for the most severe cases [15–17].

Only 10-17% of individuals with CMDs receive adequate treatment, with rates ranging from 2-4% in low- and middle-income countries to 14-22% in high-income countries [18, 19]. Contrary to clinical guidelines and patient preferences - which favor psychotherapy by a three-to-one ratio - CMDs are more frequently treated with pharmacotherapy [20–24]. Distinct factors may influence the utilization of psychotherapy and pharmacotherapy, as they have different access requirements. *Pharmacotherapy requires a medical prescription,* typically provided by general practitioners, included in reimbursement policies across many countries*, and involves less frequent follow-up. In contrast, psychotherapy requires actively seeking a specialist, scheduling regular appointments* that involve greater time and effort, often entails direct out-of-pocket payments, *and* may involve overcoming stigma associated with consulting a mental health professional [20, 25, 26].

Previous research has identified distinct factors influencing the utilization of psychotherapy and pharmacotherapy. Women, younger individuals, and those living in less rural areas were more likely to use psychotherapy, whereas older individuals, and those residing in more rural areas were more likely to use pharmacotherapy [20, 21, 23, 24, 27]. The presence of CMD may be not associated with psychotherapy use but with increased use of pharmacotherapy or combined psychotherapy and pharmacotherapy [23]. While some studies suggest that treatment utilization is equitable, others indicate potential inequalities within the health care system [23]. Specifically, higher education strongly predicted psychotherapy use, whereas individuals with lower education and retirees were more likely to receive pharmacotherapy alone [28, 29]. However, a significant gap remains, as no study has simultaneously quantified how these factors directly influence psychotherapy and pharmacotherapy initiation. Additionally, the role of mediating factors, such as CMD symptoms or perceived mental health, has not been thoroughly explored, despite their potential to yield distinct public health implications.

The Andersen’s behavioral model of health services utilization provides a robust framework for understanding how need factors (mental health status), predisposing factors (sociodemographic characteristics), and enabling factors (resources facilitating access to care) interact to influence access to care [30–32]. The COVID-19 pandemic provides an opportunity to evaluate these dynamics, as multiple cumulative stressors heightened demand for mental health care while straining healthcare systems [33]. By integrating these elements, this study seeks to offer a comprehensive analysis of the dynamics influencing treatment choices for CMDs.

Thus, this study aims to jointly investigate the factors, including pandemic-related stressors, directly and indirectly associated with the initiation of psychotherapy and pharmacotherapy in the context of CMDs, using Andersen’s conceptual model.

### 1 Theorical model

We used structural equation modeling (SEM) to assess how need, predisposing and enabling factors influence the initiation of psychotherapy or pharmacotherapy, both directly and indirectly through CMD symptom severity and self-rated mental health [34]. Based on the literature and the data collected, we formulated the following assumptions grounded in Andersen’s conceptual model (Figure 1).

**Figure 1.**
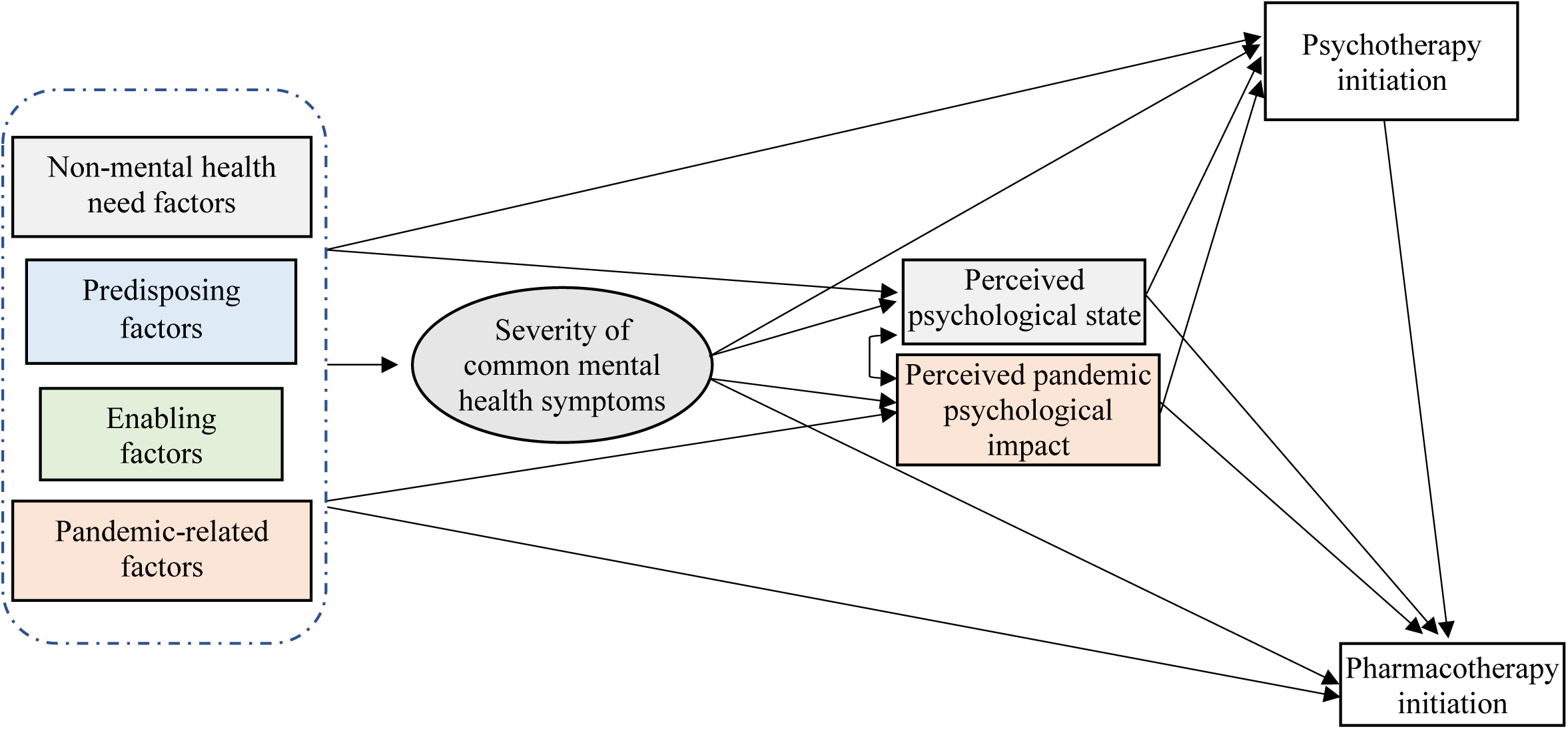
Conceptual model for the structural equation modeling: classical need (grey), predisposing (blue), and enabling (green) factors (grey), and their corresponding pandemic-related factors (red) influencing psychotherapy and pharmacotherapy initiation in the general population. Latent variables are not shown for clarity but details are presented in Supplementary material S1.

#### 1.1 Need factors

Self-rated mental health and CMD symptom intensity are strongly associated with mental health service use, though their influence may differ between psychotherapy and pharmacotherapy [23, 32]. Chronic non-psychiatric disorders and substance use disorders have been associated with higher likelihood of treatment for CMD [28, 35–37]. We hypothesize that three pandemic-related factors may increase the likelihood of seeking mental health services: i) a history of symptomatic COVID-19, as with other non-mental health disorders, and with studies suggesting SARS-CoV-2 infection alone does not predict CMD, but factors like fear and isolation may [38]; ii) heightened stress and health awareness in regions with higher COVID-19 case rates; iii) perceived mental health deterioration during the pandemic, similar to the link between self-rated mental health and treatment-seeking.

#### 1.2 Predisposing factors

Age typically follows a hill-shaped pattern, with middle-aged individuals being most likely to seek treatment, though older individuals showed a stronger association with pharmacotherapy use [27]. Individuals with lower education may be more likely to use pharmacological treatments, while those with higher education more likely to psychotherapy [29, 32]. Being married or cohabiting has been associated with lower likelihood to seek care [27, 39]. A history of adverse life events increases the likelihood of mental health care [33, 39, 40].

#### 1.3 Enabling factors

Studies have shown mixed associations between income, employment status, social support, loneliness, rural/urban residence, density of general practitioners and access to mental health care [32]. Given that the pandemic has exacerbated inequities and loneliness, we hypothesize that these factors may have influenced the use of psychotherapy and pharmacotherapy, as previously suggested [41–43]. Income loss, employment changes, and working in healthcare have been identified as enabling factors for service use during the pandemic [43]. We also considered mindfulness practices, over-the-counter (OTC) medications, and physical exercise as enabling factors, as they may alleviate CMD symptoms, potentially delaying or preventing the need for professional care, particularly when health systems are strained [44, 45].

#### 1.4 Indirect factors

We hypothesized indirect pathways thorough higher CMD symptom severity with: predisposing factors, including female sex, younger age, lower educational attainment, living alone, and history of adverse life events or stressors (e.g., job loss, divorce)[46–52]; need factors, including psychoactive substance use [53, 54], chronic non-psychiatric diseases [55, 56], potentially intensified by pandemic-related health fears [57]; enabling factors, including lower income, unemployment, low social support, loneliness, and lack of exercise [58–62], with disparities exacerbated by the COVID-19 pandemic [48–50, 63], and local deprivation and urbanicity [58, 64]; pandemic-related factors, including income loss and financial strain, self-reported COVID-19 symptoms, being a public-facing worker or living in high-risk areas [38, 62, 65, 66]. We further hypothesize that these factors contribute to perceived mental health deterioration, potentially driven by cognitive biases such as negativity bias [67, 68].

### 2 Material and methods

#### 2.1 Study design and participants

We conducted a cross-sectional study nested within the French Grippenet/Covidnet cohort in mainland France. The cohort is a voluntary online cohort established in 2012 to monitor acute respiratory infections in the general population [69]. In April 2022, the 6,944 adult participants included in this cohort were invited by email to complete a survey specific to this study on mental health care and associated factors. Information on demographic, socio-economic status, and chronic non-psychiatric comorbidities were obtained from the annual Grippenet/Covidnet inclusion survey. Other data were collected through the dedicated survey.

Participants included in this study were those who reported not using treatments prior to the pandemic, in line with the study objective of examining psychotherapy or pharmacotherapy initiated during the pandemic. Participants with extreme statistical weights were excluded to ensure robust analysis (see Statistical Analysis section for details).

Participant consent was obtained upon registration, and the study was approved by the French Advisory Committee for Research on Information Treatment in Health (authorization 11.565) and the National Commission on Informatics and Liberty (authorization DR-2012-024).

#### 2.2 Measures

##### 2.2.1 Utilization of pharmacotherapy or psychotherapy initiated during the pandemic

Pharmacotherapy utilization was assessed by asking participants: “Have you taken an antidepressant or medication for relaxation or sleep in the past month?”. Medication names provided by participants were used to verified the accuracy of the information and classify the medications as antidepressants, anxiolytics, or hypnotics based on the Anatomical Therapeutic Chemical classification system [70].

Psychotherapy utilization was assessed by asking: “Are you currently receiving regular support from a formal healthcare provider (e.g., psychiatrist, psychologist, general practitioner, or other medical doctor) intended to improve sleep, address emotional problems, or promote relaxation, excluding the use of psychotropic medications?”[71, 72]. As the present work focuses on factors associated with the initiation of psychotherapy or pharmacotherapy in the COVID-19 pandemic context, participants who reported using either of these before the pandemic were excluded from the analyses.

##### 2.2.2 Need factors

Depressive symptom severity was derived from the 9-item Patient Health Questionnaire (PHQ-9), a self-report tool assessing depressive symptom severity in the last two weeks [73]. According to this scale, scores of 0–4 indicate none or minimal depression symptoms, 5–9 mild, 10–14 moderate, 15–19 moderately severe, and 20–27 severe ones.

Anxiety symptom severity was measured using the 7-item Generalized Anxiety Disorder scale (GAD-7), which measures generalized anxiety symptoms in the last two weeks [74]. According to this scale, scores of 0–4 indicate none or minimal anxiety symptoms, 5–9 mild, 10–14 moderate, and 15–21 severe ones.

Insomnia symptom severity was assessed using the 7-item Insomnia Severity Index (ISI), which measures symptom severity in the last month [75]. According to this scale, scores of 0–7 indicate absence of insomnia, 8–14 subthreshold insomnia, 15–21 moderate, and 22–28 severe one.

Other classical need factors included self-rated mental health (good/moderate/poor) [76], chronic non-psychiatric conditions, and substance use (including tobacco, cannabidiol, cannabis, cocaine, ecstasy and other drugs) assessed as yes/no.

Pandemic-related need factors included: perceived negative psychological impact from the pandemic, a history of symptomatic COVID-19, the cumulative COVID-19 hospitalization number by department (per 100,000 inhabitants, based on government data) as a proxy for the severity of the pandemic, being a public-facing worker (defined as anyone working in direct contact with the public, including healthcare and non-healthcare workers) during the pandemic.

##### 2.2.2 Predisposing factors

Predisposing factors included age, sex, educational level (secondary education or less vs higher education), living with adult(s) (as a proxy for reduced social isolation during the pandemic), history of adverse life events, and experience of a stressful life event within the past 12 months (e.g., divorce, death of a loved one, job loss).

##### 2.2.4 Enabling factors

Individual socio-economic and psychosocial factors included: employment status, perceived financial difficulties (good/neither good nor difficult/difficult), three measures of social support (feeling supported financially, in daily life, and morally or emotionally), loneliness (4-point scale ranging from “Very lonely” to “Very surrounded”).

Three contextual factors included: the urban/rural classification (defined by the French national statistical institute, Insee, www.insee.fr), the French ecological deprivation index (Fdep, based on unemployment rate, labor rate, high school graduation rate, and median household income [77]); and the density of general practitioners per 100,000 inhabitants (defined by the French national health insurance, www.ameli.fr).

Behavioral factors included: practice of sport, practice of relaxation techniques (meditation, breathing exercises, sophrology, others), the use of over-the-counter (OTC) medications without a prescription to improve sleep, emotional balance, or aid relaxation.

The pandemic-related enabling factor was the deterioration of the financial situation during the pandemic, perceived as unchanged, improved, or degraded.

#### 2.3 Statistical methods

Missing data (2.6% of all values in the dataset) were addressed using multiple imputation by chained equations [78]. All analyses were performed on the weighted data. Details on the construction of the weights are provided in Supplementary material S2.

To perform SEM, we first tested the measurement model including two latent variables: 1) CMD symptom severity, assessed across three dimensions: depressive, anxiety, and insomnia symptoms. Given their frequent co-occurrence and shared treatment pathways, it is more appropriate to treat these symptoms as a broader construct in a non-clinical population [32, 71, 72, 79]. 2) Social support, measured across three dimensions: feeling supported financially, in daily life, and morally or emotionally [80, 81]. The correlations between the different observed variables for the same latent variable were tested using Spearman’s correlation coefficient (ρ> 0.3 for significance). To assess the unidimensionality of each latent variable, scree-plots were examined. Confirmatory Factor Analysis (CFA) was used to ensure that the observed variables contributed significantly to the corresponding latent variables. The weighted least squares means and variance (WLSMV) estimator was applied, suitable for categorical dependent variables [82]. Then, we estimated the SEM with standardized regression coefficients (from -1 to 1). The goodness of fit for the measurement and the structural model was evaluated using the following fit indices for categorical variables [83]: root mean square error of approximation (RMSEA) (desired value < 0.08), comparative fit index (CFI) (desired value > 0.95), Tucker-Lewis index (TLI) (desired value > 0.95) and standardized root mean square residual (SRMR) ≤ 0.08. Bootstrapping (1000 resamples) was applied to estimate confidence intervals for model parameters using the bias-corrected and accelerated method.

All analyses were performed in R version 4.0.2, with the ’lavaan’ package for SEM, and ’boot’ for bootstrapping. Alpha error risk was set at 5%, and all p-values were two-tailed.

## 3 Results

Of the 6,944 adults invited to participate in this study, 60.5% (n=4,199) responded. After excluding participants with extreme statistical weights (n=40) and those using treatments started before the pandemic (n=862), the final analytical sample consisted of 3,297 participants.

### 3.1 Description of study population

Table 1 summarizes the sample characteristics (raw numbers and weighted percentages). Women comprised 52.9% of the sample, 75.6% had a higher educational level, and the median age was 54 years (range: 18-91 years).

**Table 1.**
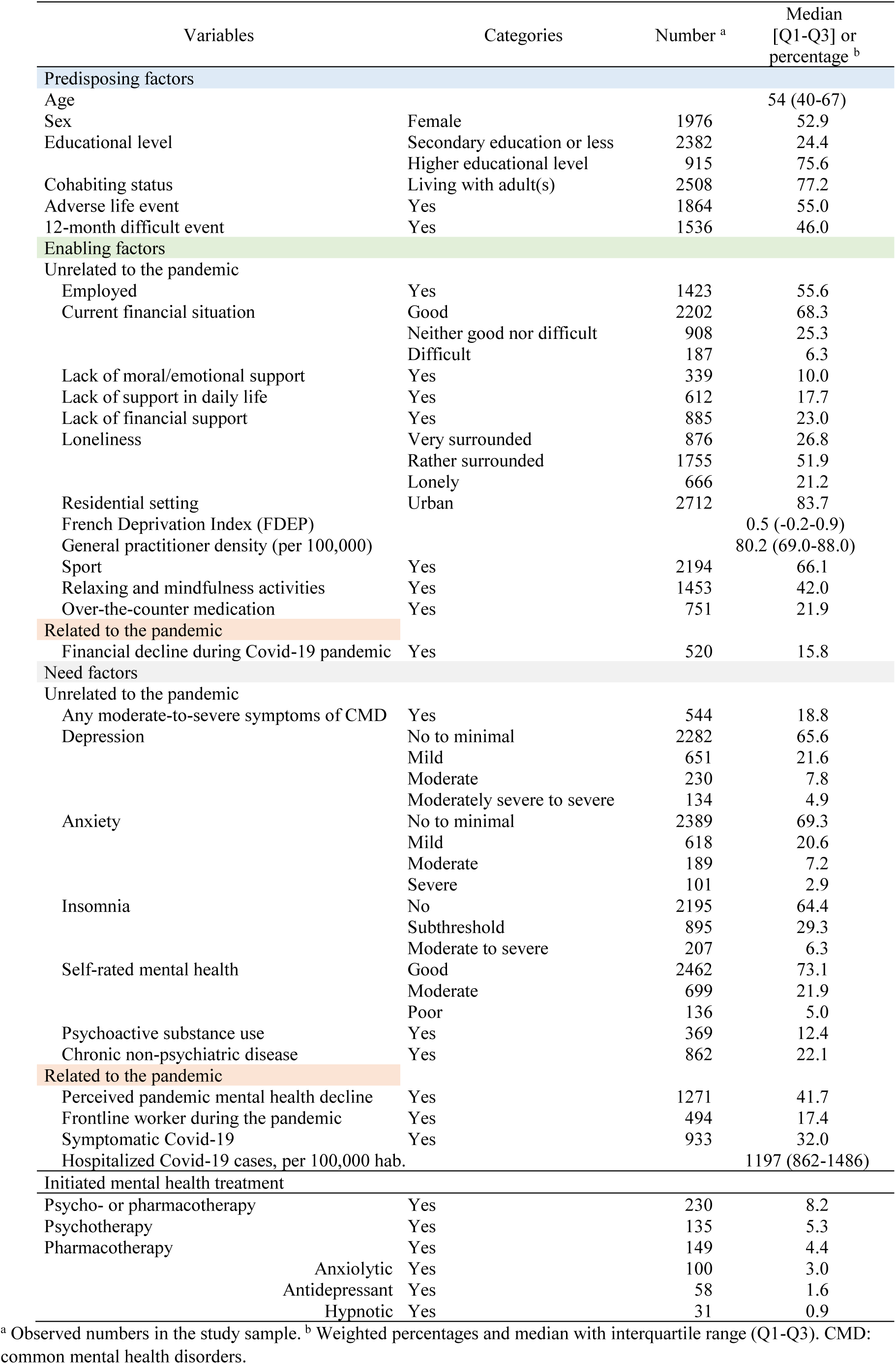
Characteristics of the study population (N=3297; weighted N = 3347).

Overall, 18.8% of participants presented moderate-to-severe symptoms, with 12.7% experiencing depressive symptoms, 10.1% anxiety symptoms, and 6.3% insomnia. Additionally, 21.9% perceived their mental health as moderate, 5.0% as poor, and 41.7% reported a negative impact of the pandemic on their mental health.

Regarding treatments, 8.2% of participants had initiated at least one during the pandemic, including 5.3% undergoing psychotherapy, and 4.4% pharmacotherapy (3.0% using anxiolytics, 1.6% antidepressants, and 0.9% hypnotics). Treatment patterns showed that 3.7% initiated only psychotherapy, 2.8% only pharmacotherapy, and 1.1% initiated both.

### 3.2 Structural equation modeling

The measurement model is provided in Supplementary material S1, and the SEM is shown in Figure 2. It demonstrated good fit indices (CFI = 0.864, TLI = 0.956, RMSEA = 0.045, SRMR = 0.040), indicating that the data aligned reasonably well with the theoretical model. The standardized estimates and their 95% confidence intervals for paths, ranked by strength of association for each outcome, are presented in Supplementary material S3. Overall, need factors had the highest effect size in both directly and indirectly influencing treatment initiation, followed by the indirect effects of loneliness and age on CMD symptom severity.

**Figure 2.**
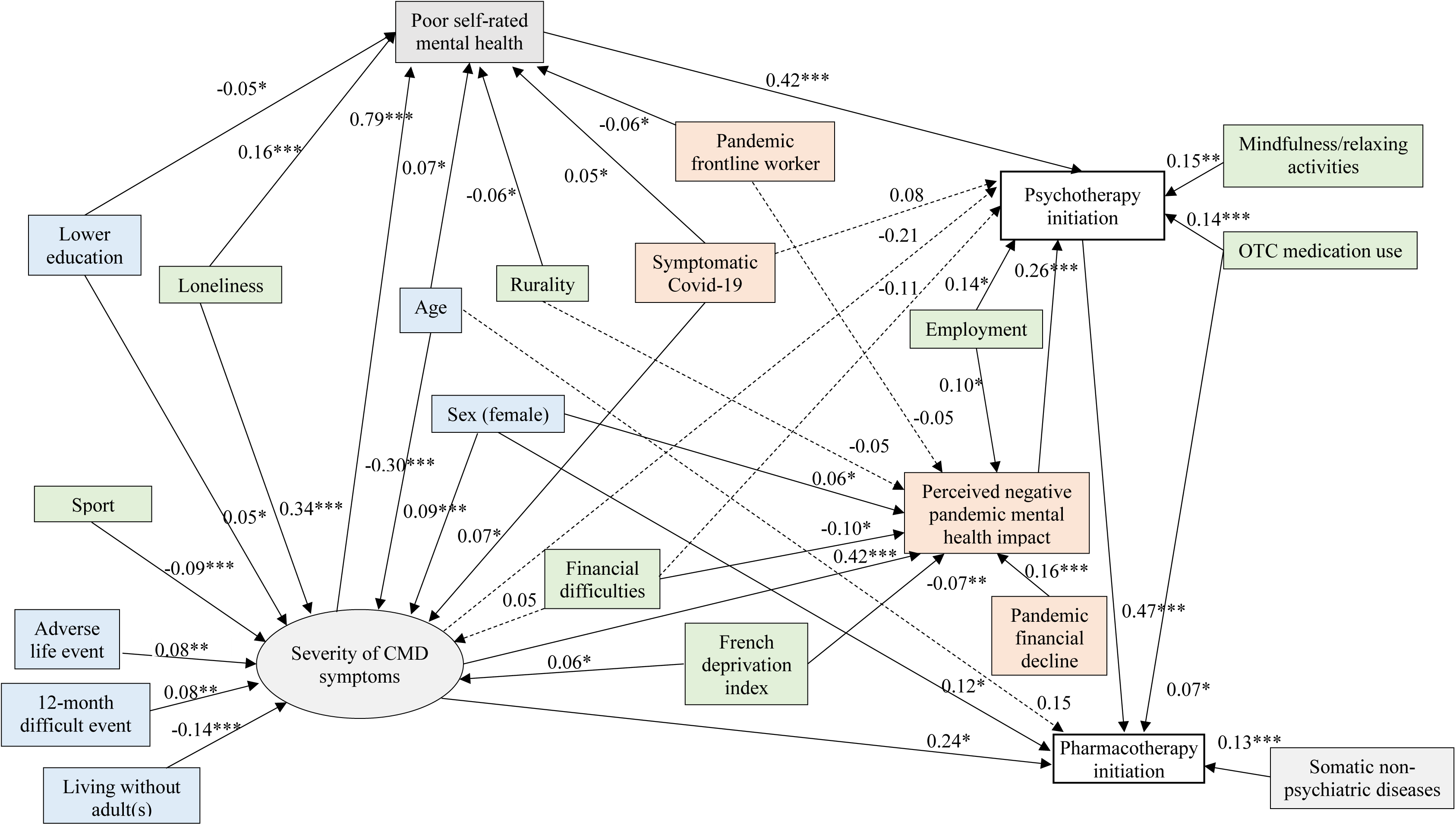
Structural equation model included need (grey), predisposing (blue), enabling (green), and pandemic-related factors (red). CMD: Common mental disorders. Solid arrow = significant, dashed arrow = borderline significant (p < 0.10). Standardized coefficients are shown with significance levels: p<0.10, *p<0.05, **p<0.01, ***p<0.001

Psychotherapy initiation was directly associated with perceived needs, including poorer self-rated mental health (β=0.42, 95%CI= [0.21, 0.63]) and a negative perceived impact of the pandemic on mental health (0.26 [0.16, 0.37]), and several enabling factors: being employed (0.14, [0.02, 0.25]), use of mindfulness or relaxing activities (0.15 [0.05, 0.24]), or of OTC medication (0.14 [0.06, 0.22]).

Pharmacotherapy initiation was directly associated with assessed needs (0.47, [0.38, 0.55]), including higher level of CMD symptoms (0.24, [0.03, 0.44]), non-psychiatric chronic disease (0.13 [0.05, 0.20]), one predisposing factors, being female (0.12 [0.01, 0.22]), and one enabling factors, the use of OTC medication (0.07 [0.00, 0.14]).

Several predisposing factors influenced treatment initiation indirectly, primarily through higher CMD symptom severity (ordered by decreasing effect size: younger, living with adults, being female, history of adverse life event and recent difficult event, lower educational level), but also through poorer self-rated mental health (older, higher educational level), and a negative perceived impact of the pandemic (female). Among these pathways, the strongest association was with CMD symptom severity, first with younger age, followed by living with adults.

Several enabling factors influenced treatment initiation indirectly, mostly through perceived needs, including poorer self-rated mental health (loneliness, living in more urban area) and a perceived negative impact of the pandemic (having a pandemic financial decline, employed, having fewer financial difficulties, living in less deprived area), but also through higher CMD symptom severity (loneliness, not practicing sport, living in more deprived area,). Among these, the strongest association was between CMD symptom severity and loneliness, followed by its association with poorer self-rated mental health, and the effect of financial decline on perceived pandemic impact.

## 4 Discussion

Our study extends previous research on Andersen’s behavioral model by providing a comprehensive analysis of how need, predisposing, and enabling factors, including pandemic-related factors, simultaneously contribute to the initiation of treatments for CMD. We identified distinct pathways for psychotherapy and pharmacotherapy initiation. Psychotherapy initiation was directly associated with perceived need and several enabling factors, whereas pharmacotherapy initiation was directly linked to evaluated need, one predisposing factors, and one enabling factors. Indirectly, predisposing factors influenced treatment initiation primarily through CMD symptoms, while enabling and pandemic-related factors exerted their effects mainly through poorer perceived need, but also through CMD symptoms.

Distinct need factors predominantly shape treatment pathways for CMD: psychotherapy is driven by perceived need, while pharmacotherapy is shaped by assessed clinical need. This central role of need factors aligns with previous research, both before and during the pandemic [32, 39, 43, 84–87]. Since pharmacotherapy follows contact with a healthcare professional, perception plays a critical role in initiating both types of treatment. This finding is in line with results obtained in countries with different income levels, where low perceived need and attitudinal barriers have been identified as greater obstacles to mental health service use, rather than structural barriers [32, 88]. Strengthening mental health literacy may therefore help reduce treatment gaps [87]. The near-significant link between symptomatic COVID-19 and psychotherapy initiation suggests that responses to infectious disease may act as drivers of help-seeking, aligning with previous research [38, 89]. In contrast, the direct association between CMD symptoms and pharmacotherapy highlights the role of clinical assessment once individuals engage with the healthcare system. Similarly, the association between chronic non-psychiatric conditions and pharmacotherapy, consistent with prior studies [32, 35], may reflect the fact that individuals with ongoing care relationships are more likely to receive medication, whether due to patient preference for familiar interventions or physicians’ prescribing habits.

While sex has a direct role in pharmacotherapy initiation, other predisposing factors mainly influence treatment initiation earlier in the pathway, through CMD symptom severity and perceived mental health. The association between women and pharmacotherapy is consistent with previous research indicating that women are more likely than men to use anxiolytics for acute social and emotional issues [90]. The lack of a direct association with psychotherapy initiation contrasts with pre-pandemic studies linking sex to mental health help-seeking [27, 32, 33, 39, 40]. This discrepancy may reflect pandemic-specific shifts in help-seeking dynamics, as reported by others [84], or be due to confounding factors adjusted for in our analysis [35, 90]. Regarding indirect role of other predisposing factors, two findings are noteworthy due to their contrasting associations. Lower education and younger age were both positively associated with increased CMD symptom severity, but negatively associated with poorer self-rated mental health. These contrasts may highlight a need to improve mental health literacy among individuals with lower education levels and younger populations.

We showed that enabling factors influence various stages of the pathway to CMD treatment initiation. Although their overall effect size was small, aligning with pre-pandemic studies [32, 87, 91], enabling factors may have played a more substantial role in disadvantaged populations or earlier in the pandemic, with their impact diminishing by 2022. This limited influence may also reflect the equalizing effect of the French healthcare system, which reduces access disparities, particularly for general practice and pharmacotherapy. Nonetheless, their importance remains due to their modifiability within Andersen’s model [30, 31]. Loneliness was the most strongly associated enabling factor, albeit indirectly, underscoring its key role in CMD symptom severity and self-rated mental health, as highlighted in both pandemic and pre-pandemic studies [60, 92–94]. Employment was positively associated with psychotherapy initiation, and fewer financial difficulties showed a near-significant positive association, suggesting inequities in access to psychotherapy but not pharmacotherapy, consistent with the lack of reimbursement for psychological support in France at the time. Mindfulness activities and OTC medication were positively linked to treatment initiation, indicating that such self-care strategies may complement, but not substitute, formal support. Practicing sport was negatively associated with CMD symptom severity, reinforcing its protective role. The relationship between enabling factors and perceived need is noteworthy. Disadvantaged individuals (those in deprived areas or facing financial hardship) were less likely to report a negative mental health from the pandemic, potentially reflecting resilience, stigma, or lower mental health literacy. Conversely, being employed and experiencing income loss during the pandemic were more likely to report a perceived negative mental health impact, possibly reflecting greater vulnerability among those more affected by pandemic-related disruptions. Urban living was linked to poorer self-rated mental health, potentially due to increased stressors, better awareness of services or less stigma compared to rural areas.

This study is strengthened by the use of SEM to assess direct and indirect pathways to psychotherapy and pharmacotherapy, but several limitations should be noted. First, the cross-sectional design precludes causal inferences. Second, despite statistical weights, the voluntary, non-random sample may limit representativeness, particularly for youth and disadvantaged populations. Third, self-reported data and the absence of pre-pandemic baseline may introduce recall bias. Fourth, we did not capture participants who initiated treatment during the pandemic but later discontinued it. However, the two-year data still reflect sustained treatment use, as the average help-seeking delay is typically measured in years [95–97]. Fifth, the analysis focused only on the use of pharmacotherapy and psychotherapy, without details on treatment types or providers, and on mindfulness and relaxation activities, without knowing if they were used for self-management or with professional guidance. Sixth, we did not assess the functional impact of CMD symptoms or quality of life, which are potentially crucial factors in treatment decisions. Finally, confounding factors may not have been fully accounted for in the analysis.

## 5 Conclusion

This study identified distinct pathways to psychotherapy and pharmacotherapy initiation for CMDs, offering insights to improve access to both treatments. Strengthening mental health literacy, particularly among disadvantaged groups, and promoting self-management strategies, such as mindfulness may help address treatment gaps. Additionally, alleviating structural barriers for psychotherapy is essential for equitable access to mental health care. Further research should evaluate the effectiveness of these interventions in improving mental health service utilization.

## Conflicts of interest

The author(s) declare none.

## Funding

This research received no specific grant from any funding agency, commercial or not-for-profit sector. The study was conducted within the authors’ regular salaried positions.

## Author’s contribution

All authors participated in the design of the study. MP conducted the analyses. MP wrote the first draft of this report. All authors contributed to the review of the manuscript, read the manuscript and approved the final version.

## Supplementary material

S1. Measurement model

S2. Weighted data

S3. Structural equation model: standardized estimates, confidence intervals, and p-values for need (grey), predisposing (blue), enabling (green), and pandemic-related factors (red), ranked by association strength for each outcome.

## Data Availability

All data produced in the present study are available upon reasonable request to the authors

## Notes

### Competing Interest Statement

The authors have declared no competing interest.

### Funding Statement

This study did not receive any funding

### Author Declarations

The study was approved by the French Advisory Committee for Research on Information Treatment in Health (authorization 11.565) and the National Commission on Informatics and Liberty (authorization DR-2012-024). The surveys used in and performed for our study were de-identified

